# Uncovering Genetic Risk Beyond Diagnoses in Suicidal Thoughts and Behaviors: Insights from *All of Us*

**DOI:** 10.1101/2025.04.18.25326075

**Authors:** Phil H. Lee, Brandon T. Sanzo, Younga H. Lee, Daniel H. Jung, Jodi M. Gilman, Matthew K. Nock, Jordan W. Smoller, Richard T. Liu, Ronald C. Kessler

## Abstract

**Importance:** Suicide is a leading cause of death worldwide, yet risk prediction remains imprecise. While psychiatric disorders are strongly associated with suicide-related outcomes, most individuals with these conditions never exhibit suicidal behaviors. Polygenic risk scores (PRSs) may help identify additional vulnerability factors beyond clinical diagnoses.

**Objective:** To evaluate the independent and interactive effects of polygenic risk for psychiatric disorders and clinical diagnoses on suicidal ideation (SI) and suicide attempts (SA) in a large, ancestrally diverse cohort.

**Design:** Cross-sectional analysis of genetic and survey data from the *All of Us* Research Program.

**Setting:** Population-based cohort study leveraging a diverse U.S. sample.

**Participants:** 41,379 adults with genetic data and self-reported psychiatric diagnoses, SI, and SA.

**Main Outcomes and Measures:** Lifetime SI and SA, assessed via self-reported surveys. Predictors included lifetime psychiatric diagnoses on 13 categories and PRSs for depression, bipolar disorder, and PTSD, derived from multi-ancestry genome-wide association studies. Ancestry-stratified multinomial logistic regression analyses were performed for African, Admixed Hispanic/Latino, and European American groups, followed by fixed-effects meta-analysis, adjusting for age, sex at birth, and socioeconomic factors.

**Results:** Among 41,379 participants, 28.5% reported SI, and 12.6% reported SA. All psychiatric disorders were significantly associated with both outcomes, with depression, bipolar disorder, and PTSD showing the strongest independent effects (ORs=2.81-7.73 for SA, 1.62-3.32 for SI, all FDR < 0.05). Each additional psychiatric diagnosis more than doubled the odds of SA (OR=2.16 95% CI: 2.10-2.21). PRSs for depression, bipolar disorder, and PTSD remained significantly associated with SI and SA after adjusting for clinical diagnoses and sociodemographic covariates. For SA, depression PRS showed the strongest association (OR=1.36 [1.30–1.41], p=1.42×10^-55^), followed by PTSD (OR=1.33 [1.28-1.39], p=6.91×10^-45^) and bipolar disorder (OR=1.18 [1.13-1.23], p=1.41×10^-16^). Effect sizes were comparable among individuals with and without clinical diagnoses, suggesting transdiagnostic relevance.

**Conclusions:** Polygenic risk for psychiatric disorders showed modest but significant associations with SI and SA, independent of clinical diagnoses and sociodemographic factors. These findings highlight the value of genetic information in identifying vulnerability not fully captured by diagnostic categories and underscore the importance of multi-dimensional approaches to suicide risk assessment across diverse populations.

**KEY POINTS:** *Question:* Do polygenic risk scores (PRS) for psychiatric disorders independently predict suicidal ideation (SI) and suicide attempts (SA) beyond clinical diagnoses?

*Findings:* In 41,379 *All of Us* participants, socioeconomic adversity and psychiatric diagnoses were strongly associated with SI and SA. PRSs for depression, bipolar disorder, and post-traumatic stress disorder (PTSD) showed significant and independent associations with SI and SA. These associations remained regardless of clinical diagnoses, suggesting genetic risk reflects vulnerability not fully captured by diagnostic categories.

*Meaning:* While PRSs have limited predictive value individually, integrating genetic, clinical, and socioeconomic factors may enhance understanding of suicide risk and improve risk assessment.

## INTRODUCTION

Suicide represents a global public health emergency, responsible for over 720,000 lives annually,^1^ including more than 49,000 in the United States (US) in 2022.^2^ Many more people experience suicidal ideation (SI) and suicide attempts (SA), with profound impacts on individuals, families, and communities.^3^

Psychiatric disorders are among the strongest predictors of suicide-related outcomes,^4^ with 80– 95% of individuals who die by suicide having a mental health condition.^5^ Major depressive disorder, bipolar disorder, substance use disorders, and post-traumatic stress disorder (PTSD) are particularly high-risk.^6^ However, most individuals with psychiatric diagnoses do not engage in suicidal behaviors,^4^ underscoring the critical need for more refined risk indicators to explain differential outcomes.

The stress-diathesis model^7^ suggests that suicide risk arises from both stressors—such as psychiatric illness—and underlying vulnerabilities, including genetic predisposition.^8,9^ Recent genome-wide association studies (GWASs) have identified both shared and unique genetic contributions to psychiatric disorders and suicidal behaviors.^10–15^ Polygenic risk scores (PRSs),^16^ which aggregate genetic liability based on these GWAS findings, offer a promising avenue to quantify inherited vulnerability. Indeed, individual psychiatric disorder PRSs—particularly for depression—have been robustly associated with suicide-related outcomes, encompassing SI, SA, and suicide death.^17–22^

Despite these advances, few studies have examined whether genetic risk adds independent value *beyond* psychiatric diagnoses. In a notable example, Stein et al.^21,22^ demonstrated that depression PRS was associated with SA independent of personal and family history of depression in a US military sample, suggesting that PRSs may capture transdiagnostic genetic liability. Whether these findings generalize to more diverse, population-based cohorts and across multiple psychiatric disorders and broader sociodemographic contexts remains an open question.

Moreover, the vast majority of PRS research has been conducted in individuals of European ancestry,^22–27^ limiting clinical relevance across diverse populations. Although recent studies have begun to include multi-ancestry cohorts,^17,28^ findings are often inconsistent, and research in large, ancestrally diverse, population-based samples remains limited—highlighting an urgent need for more inclusive approaches to genomic research and precision psychiatry.

To address these gaps, we leverage data from the *All of Us* Research Program,^29–31^ a large, nationwide U.S. cohort enriched with extensive genetic, clinical, and sociodemographic information. Our study aims to: (1) characterize sociodemographic differences in SI, SA, and non-suicidal individuals; (2) evaluate associations between psychiatric disorders and suicide-related outcomes; (3) assess the independent value of PRSs for psychiatric disorders beyond sociodemographic factors and clinical diagnoses; and (4) test PRS-by-diagnosis interactions to identify high-risk subgroups. By integrating genetic, clinical, and sociodemographic factors in a diverse population, our study seeks to advance more equitable and refined suicide prevention strategies.

## METHODS

### Study Cohort

The *All of Us* Research Program^29–31^ is an ongoing, nationwide biomedical initiative designed to advance precision medicine by recruiting a demographically diverse cohort of one million participants across the US. This study utilized data from the curated release v8 (Controlled Tier 2024Q3R5), which includes participant data collected through October 1, 2023. All analyses were conducted between June 2024 and April 2025 using the *All of Us* Researcher Workbench, a secure cloud-based platform available to authorized investigators.

### Ethics Review

The *All of Us* data collection was conducted under centralized Institutional Review Board (IRB) approval, with informed consent obtained from all participants. This study adhered to ethical guidelines outlined in the *All of Us* Code of Conduct. The Massachusetts General Hospital IRB reviewed the study protocol and determined that this secondary analysis of de-identified *All of Us* data is exempt from human subjects research.

### Suicide-Related Outcomes

SI and SA were defined using self-reported survey items from the Emotional Health and Well-Being Survey. SI was assessed using the question: *“Did you ever in your life have thoughts of killing yourself?”.* SA was determined by the question: *“Did you ever make a suicide attempt where you purposefully hurt yourself with at least some intention to die?”* Individuals who selected *skip or refused to answer* were excluded from the analysis (N=572, 0.59%). Participants were categorized into three groups: those with SA, those with SI only, and those without any suicide-related outcomes (controls) (**eFigure 1**).

### Lifetime Diagnoses of Psychiatric Disorders and Comorbidity

In the Personal and Family Health History survey, participants reported lifetime diagnoses of 13 mental health or substance use disorders: alcohol use disorder, anxiety reaction/panic disorder, attention-deficit/hyperactivity disorder (ADHD), autism spectrum disorder, bipolar disorder, depression, drug use disorder, eating disorder, personality disorder, PTSD, schizophrenia, social phobia, and other mental health conditions. Psychiatric comorbidity, representing the cumulative burden of psychiatric conditions, was measured as the total count of distinct lifetime diagnoses reported by each participant, ranging from 0 to 13.

### Sociodemographic Factors

Sociodemographic data were obtained from the Basics survey, including age, sex at birth, education, employment status, health insurance, income, marital status, self-identified race/ethnicity, and sexual orientation (**eTable 1**).

### Polygenic Risk of Psychiatric Disorders

Genome sequencing, quality control (QC), and ancestry predictions were conducted centrally by the *All of Us* Research team and are detailed elsewhere.^32^ PRSs were computed using PRS-CS^33^ and PLINK.^34^ To ensure broad applicability across the diverse *All of Us* cohort, we constructed PRSs for psychiatric disorders most robustly associated with suicidal behaviors in previous studies and having well-powered, multi-ancestry GWAS summary statistics: major depression (N = 688,808 cases, 4.36 million controls),^35^ bipolar disorder (N = 158,036 cases, 2.8 million controls),^36^ and PTSD (N = 150,760 cases, 1.28 million controls)^37^ (**eTable 2**). PRSs were calculated separately for participants of African/African American (AFR), Admixed Hispanic/Latino American (AMR), and European (EUR) ancestry to account for population structure and ensure appropriate linkage disequilibrium (LD) modeling. Analyses were adjusted for ancestry-related genetic principal components (PCs) to minimize population stratification bias.

### Statistical Analysis

We performed descriptive analyses to characterize sociodemographic and psychiatric profiles across SI, SA, and control groups (*Kruskal-Wallis* tests for continuous variables and *chi-square* tests for categorical variables). Multinomial logistic regression was used to examine associations between risk variables (socioeconomic factors, psychiatric diagnoses, and PRSs) and suicide-related outcomes (SI, SA, and controls), with the control group as the reference. Models were adjusted for age, self-reported sex at birth, and genetic PCs. The proportion of variance explained by each predictor was assessed using *McFadden*’s *pseudo R*^2^. To evaluate the added predictive utility of PRSs, we compared models with and without a PRS using likelihood ratio tests and changes in *McFadden*’s *pseudo R*^2^. We also tested interactions between PRSs and psychiatric diagnoses to identify subgroups at elevated risk. All analyses were stratified by genetic ancestry (European, African, and Hispanic/Latino) and meta-analyzed using fixed-effects models. Analyses were conducted in R (v4.3.1) with multiple comparisons controlled using the false discovery rate (FDR).

### Sensitivity Analyses

To ensure robustness, we conducted multiple analyses: (1) sequential adjustment models (minimal, socioeconomic, and psychiatric diagnoses); (2) assessment of psychiatric comorbidity’s influence on PRS associations; (3) varying case and control group definitions (exclusive/non-exclusive SI, with/without psychiatric disorders); (4) alternative statistical methods (ordinal regression); and (5) different PRS construction approaches. Detailed methods are provided in the Supplementary Materials.

## RESULTS

### Study Cohort

This cross-sectional study analyzed data from 41,379 individuals in the *All of Us* Research Program, integrating self-reported suicide-related outcomes, psychiatric diagnoses, and sequencing data passing QC (**eFigure 1**). Among these, 24,398 participants (59.0%) reported no suicide-related outcomes, 11,773 (28.5%) reported SI only, and 5,208 (12.6%) reported SA.

**Table 1** summarizes sociodemographic characteristics across these groups. Those with a history of SA were the youngest on average (52.76 ± 15.03 years), followed by those reporting SI only (57.04 ± 15.99 years), while controls were oldest (61.88 ± 14.70 years, *Kruskal-Wallis* test p < 0.001). Participants self-identifying as female at birth were disproportionately represented in SA (78.9%) compared to SI (69.0%) and control (70.2%) groups (chi-square p < 0.001). Racial disparities were notable, with individuals identifying as multiracial being overrepresented in the SA group (8.9% vs. 3.9% in controls, chi-square p-value < 0.001).

**Table 1.** Sociodemographic Characteristics of Study Participants Stratified by Suicide-Related Outcomes in the *All of Us* Research Program (N=41,379). Group differences in sociodemographic characteristics were assessed using Kruskal-Wallis tests for continuous variables and chi-square tests for categorical variables. Missing includes the number of participants who chose skip, refuse to answer, or not applicable as a response. All responses are based on self-report surveys conducted by study participants. Following the *All of Us* of Research policy, we excluded groups with less than 20 participants from this table.

| Characteristics | Responses | Controls | Suicidal Ideation Only | Suicide Attempts | P value |
| --- | --- | --- | --- | --- | --- |
|  |  | N=24398 (59%) | N=11773 (28.5%) | N=5208 (12.6%) |  |
| Age | Age (years) mean ± SD | 61.88 (±14.70) | 57.04 (±15.99) | 52.76 (±15.03) | $p < 1 \times 10^{-16}$ |
| Sex Assigned at Birth | Female | 17125 (70.2%) | 8126 (69.0%) | 4111 (78.9%) | $p < 1 \times 10^{-16}$ |
|  | Male | 7151 (29.3%) | 3599 (30.6%) | 1072 (20.6%) |  |
|  | Other | 122 (0.5%) | 48 (0.4%) | 25 (0.5%) |  |
| Self-Reported Race/Ethnicity | Black or African American | 1048 (4.4%) | 345 (3.0%) | 292 (5.7%) | $p < 1 \times 10^{-16}$ |
|  | Hispanic or Latino | 1092 (4.5%) | 415 (3.6%) | 246 (4.8%) |  |
|  | More than One Population | 944 (3.9%) | 593 (5.1%) | 452 (8.9%) |  |
|  | White | 20416 (85.0%) | 9991 (86.4%) | 4003 (78.4%) |  |
|  | Missing | 503 (2.1%) | 211 (1.8%) | 112 (2.2%) |  |
| Highest Level of Education | Advanced Degree | 9613 (39.6%) | 4456 (38.0%) | 1145 (22.4%) | $p < 1 \times 10^{-16}$ |
|  | College Graduate | 7306 (30.1%) | 3697 (31.6%) | 1399 (27.3%) |  |
|  | College (One to Three Years) | 5457 (22.5%) | 2785 (23.8%) | 1922 (37.5%) |  |
|  | Missing | 1835 (7.6%) | 765 (6.5%) | 634 (12.4%) |  |
| Employment Status | Yes | 12299 (50.4%) | 6524 (55.4%) | 2562 (49.2%) | $p < 1 \times 10^{-16}$ |
|  | No | 11939 (48.9%) | 5179 (44.0%) | 2606 (50.0%) |  |
|  | Missing | 160 (0.7%) | 70 (0.6%) | 40 (0.8%) |  |
| Homeownership Status | Yes | 18370 (75.3%) | 7755 (65.9%) | 2513 (48.3%) | $p < 1 \times 10^{-16}$ |
|  | No | 5811 (23.8%) | 3929 (33.4%) | 2639 (50.7%) |  |
|  | Missing | 217 (0.9%) | 89 (0.8%) | 56 (1.1%) |  |
| Poverty<br>(Annual Income less<br>than 35K) | Yes | 3477 (14.3%) | 2186 (18.6%) | 1945 (37.3%) | $p < 1 \times 10^{-16}$ |
|  | No | 19030 (78.0%) | 8902 (75.6%) | 2933 (56.3%) |  |
|  | Missing | 1891 (7.8%) | 685 (5.8%) | 330 (6.3%) |  |
| Health Insurance<br>Ownership Status | Yes | 23887 (97.9%) | 11485 (97.6%) | 4970 (95.4%) | $p < 1 \times 10^{-16}$ |
|  | No | 336 (1.4%) | 227 (1.9%) | 188 (3.6%) |  |
|  | Missing | 175 (0.7%) | 61 (0.5%) | 50 (1.0%) |  |
| Having Partner | Yes | 16283 (66.7%) | 7089 (60.2%) | 2525 (48.5%) | $p < 1 \times 10^{-16}$ |
|  | No | 7968 (32.7%) | 4614 (39.2%) | 2633 (50.6%) |  |
|  | Missing | 147 (0.6%) | 70 (0.6%) | 50 (1.0%) |  |
| Heterosexual<br>Orientation | Yes | 22333 (91.5%) | 9509 (80.8%) | 3632 (69.7%) | $p < 1 \times 10^{-16}$ |
|  | No | 1874 (7.7%) | 2165 (18.4%) | 1522 (29.2%) |  |
|  | Missing | 191 (0.8%) | 99 (0.8%) | 54 (1.0%) |  |

Individuals experiencing socioeconomic disadvantages—including financial hardship, lower educational attainment, lack of partnership, housing instability, sexual minority status, and a lack of health insurance coverage—generally exhibited a gradient of risk, with those who had SA experiencing the highest levels of disadvantage, followed by individuals with SI, and then controls (**eTable 3**). These disparities were significant across all measures we examined, reinforcing the strong link between socioeconomic adversity and suicide risk (all p<0.001).

### Associations Between Psychiatric Disorders and Suicide-Related Outcomes

All 13 psychiatric disorders examined were strongly associated with increased risk of SI and SA (**Table 2, eTable 4**). Across all disorders, we observed a consistent gradient of prevalence estimates: highest among SA, intermediate in those with SI only, and lowest in controls (all chi-square test p<0.001).

**Table 2.** Prevalence of psychiatric disorders among the groups of participants stratified by suicide-related outcomes in the *All of Us* Research Program (N=41,379). All false discovery rate (FDR) < 0.001. ADHD = attention deficit/hyperactivity disorder; PTSD = post-traumatic stress disorder; SA = suicide attempts; SI = suicidal ideation only.

| Psychiatric Disorders | Controls (%) | SI (%) | SA (%) | Chi Square P |
| --- | --- | --- | --- | --- |
| Alcohol use disorder | 4.35 | 7.9 | 13.79 | $p < 1 \times 10^{-16}$ |
| Anxiety reaction/panic disorder | 25.22 | 41.31 | 57.99 | $p < 1 \times 10^{-16}$ |
| ADHD | 7.25 | 13.77 | 20.78 | $p < 1 \times 10^{-16}$ |
| Autism spectrum disorders | 0.64 | 1.79 | 4.05 | $p < 1 \times 10^{-16}$ |
| Bipolar disorder | 2.11 | 5.81 | 20.28 | $p < 1 \times 10^{-16}$ |
| Depression | 34.11 | 64.78 | 80.91 | $p < 1 \times 10^{-16}$ |
| Drug use disorder | 1.6 | 2.77 | 9.39 | $p < 1 \times 10^{-16}$ |
| Eating disorder | 3.47 | 7.82 | 15.25 | $p < 1 \times 10^{-16}$ |
| Personality disorder | 0.57 | 1.9 | 9.58 | $p < 1 \times 10^{-16}$ |
| PTSD | 7.66 | 18.16 | 40.02 | $p < 1 \times 10^{-16}$ |
| Schizophrenia | 0.22 | 0.42 | 2.15 | $p < 1 \times 10^{-16}$ |
| Social phobia | 1.99 | 5.67 | 11.67 | $p < 1 \times 10^{-16}$ |
| Other mental disorders | 1.43 | 3.27 | 6.45 | $p < 1 \times 10^{-16}$ |
| No Diagnosis | 47.02 | 19.69 | 7.24 | $p < 1 \times 10^{-16}$ |
| Any Diagnoses | 52.98 | 80.31 | 92.76 | $p < 1 \times 10^{-16}$ |
| Two or More Diagnoses | 22.9 | 49.45 | 74.33 | $p < 1 \times 10^{-16}$ |

Depression was most common among those with suicide-related behaviors, affecting 80.91% of the SA group, 64.78% of those with SI only, and 34.11% of controls. Anxiety disorders followed a similar pattern (57.99%, 41.31%, and 25.22%, respectively). Notably, certain disorders showed particularly marked increase in prevalence in the SA group, including PTSD (40.02% vs. 7.66% in controls) and bipolar disorder (20.28% vs. 2.11% in controls).

The burden of psychiatric comorbidity increased substantially across groups (**eTable 5**). While 52.98% of controls reported a lifetime diagnosis of at least one psychiatric disorder, this rose to 80.31% in those with SI and 92.76% in SA. Multiple disorders (two or more) were present in 74.33% of SA compared to 49.45% of individuals with ideation only and 22.90% of controls.

To quantify these associations while accounting for potential confounders, we performed multinomial logistic regression analyses adjusting for age, sex, and various socioeconomic factors (**Figure 1, eTable 6, eFigure 2**). When examined individually, personality disorder exhibited the strongest association with SA (adjusted Odds Ratio (OR) = 10.18 [8.19–12.64], R^2^=0.81%, p < 0.001), followed by bipolar disorder (OR = 7.73 [6.83-8.75], R^2^=1.59%, p < 0.001) and depression (OR = 6.41 [5.91-6.94], R^2^=5.46%, p < 0.001). The presence of any psychiatric disorders increased the odds of reporting SA more than eightfold (OR = 8.38 [7.45-9.43], R^2^=4.61%, p < 0.001).

**Figure 1.**
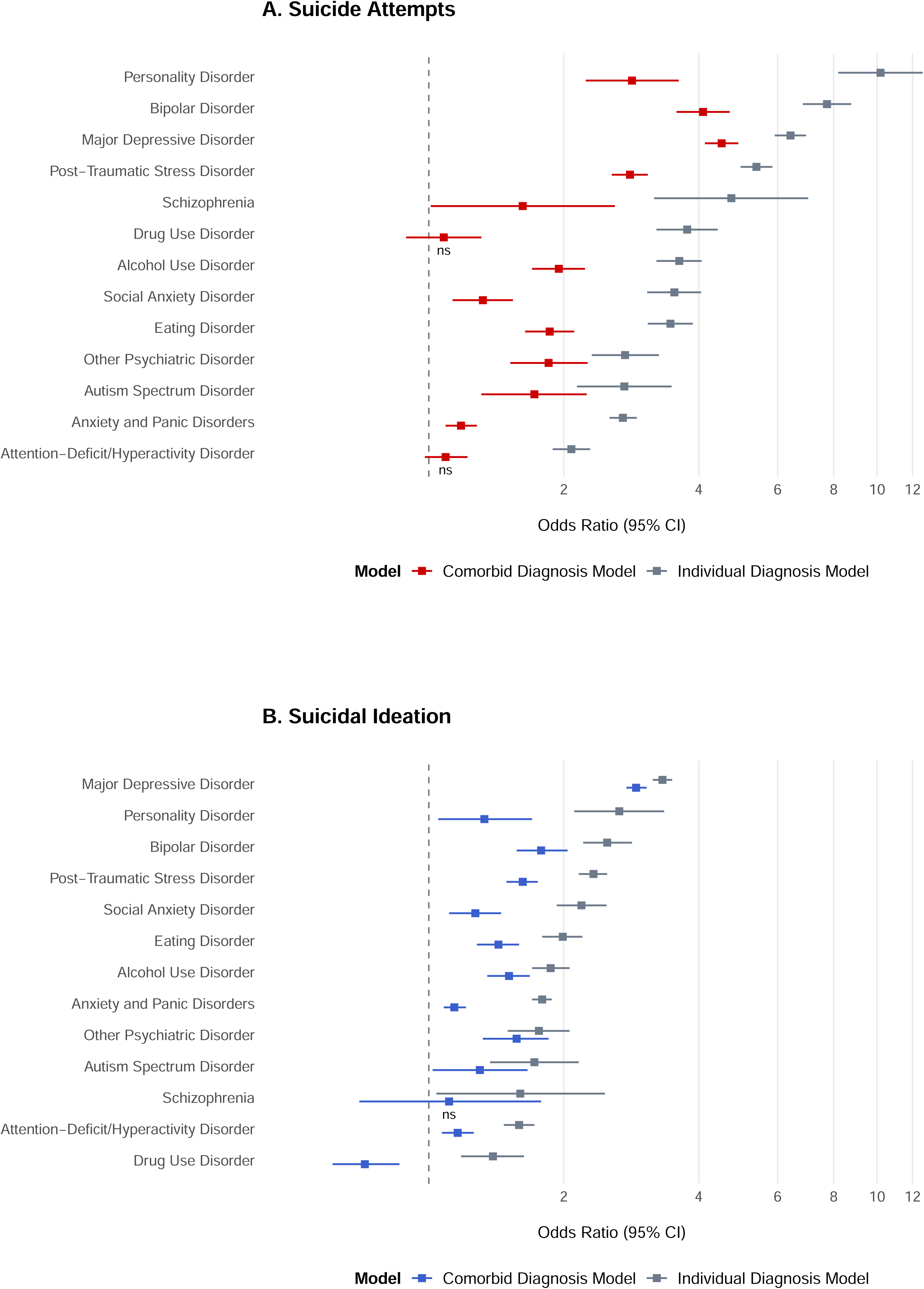
Associations Between Psychiatric Disorders and Suicide-Related Outcomes. Plots depicting adjusted odds ratios (ORs) and 95% confidence intervals (CIs) for the associations between psychiatric disorders and suicide-related outcomes. For clarity, ORs are displayed on a log scale. Panel A presents results for suicide attempts (SA), while Panel B presents results for suicidal ideation (SI). The *Individual Diagnosis Model* (gray) represents associations when each disorder is examined separately, adjusting for age, sex, and five socioeconomic factors (employment status, having partner, health insurance ownership, poverty (annual income less than $35K), and minority sexual orientation). The *Comorbid Diagnosis Model* (red/blue) includes all psychiatric disorders simultaneously in the same regression model, with the same covariates included in the Individual Diagnosis Model. Effects that are not significant after multiple testing correction at FDR < 5% are marked with “ns” (non-significant) on the plot. Complete statistical data are provided in eTables 6-7.

The impact of psychiatric comorbidity was particularly notable, with each psychiatric disorder diagnosis more than doubling the odds of SA (OR=2.16 [2.10-2.21], R^2^=6.73%) compared to controls. Individuals diagnosed with two or more psychiatric disorders showed significantly higher odds of SA (OR = 3.83 [3.51-4.17], p < 0.001) compared to those with a single diagnosis, suggesting that cumulative psychiatric burden is associated with suicide risk beyond the effects of individual disorders alone.

Notably, even when all psychiatric disorders are included in the same model (**eTable 7**), most disorders still maintained independent and significant associations with suicide-related outcomes (Comorbid Diagnosis Model in **Figure 1**). For SA, depression showed the strongest independent association (OR=4.5 [4.13-4.9]), followed by bipolar disorder (OR=4.09 [3.57-4.69]), personality disorders (OR=2.84 [2.24-3.61]), and PTSD (OR=2.81 [2.56-3.08]). For SI, depression similarly exhibited the strongest association (OR=2.9 [2.76-3.06]), followed by bipolar disorder (OR=1.78 [1.57-2.04]) and PTSD (OR=1.62 [1.49-1.75]). After full adjustment, ADHD and drug use disorder retained significant associations with SI but not with SA.

### Independent Contribution of Polygenic Risk Scores for Suicide-Related Outcomes

Next, we examined whether genetic risk, as captured by PRSs, contributes independently to predicting suicide risk beyond clinical diagnoses and sociodemographic factors. We focused on PRSs for depression, bipolar disorder, and PTSD, as these disorders showed the strongest independent associations with SI and SA (**eTable 7**). Additionally, these PRSs were derived from well-powered, multi-ancestry GWAS datasets,^35–37^ ensuring broad applicability across the diverse *All of Us* sample.

Figure 2 summarizes the associations of these PRSs with suicide-related outcomes, adjusting for age, sex, socioeconomic factors, and corresponding psychiatric diagnoses (**eTables 8-9, eFigures 3-4**). Across all three PRSs, we observed significant, independent associations with both SI and SA. For SA, the depression PRS showed the strongest association, with each standard deviation (SD) increase in PRS associated with 1.36-fold higher odds of SA (meta-analyzed OR=1.36 [1.30-1.41], *p*=1.42×10^-55^). The PTSD PRS also showed a robust association (OR=1.33 [1.28-1.39], *p*=6.91×10^-45^), followed by the bipolar disorder PRS (OR=1.18 [1.13-1.23], *p*=1.41×10^-16^). For SI, the effect sizes were more modest but statistically significant. The depression PRS was the strongest predictor (OR=1.14 [1.11-1.17], *p*=4.78×10^-24^), followed by the bipolar disorder PRS (OR=1.13 [1.10-1.16], *p*=1.20×10^-19^) and the PTSD PRS (OR=1.13 [1.10-1.16], *p*=9.61×10^-20^).

**Figure 2.**
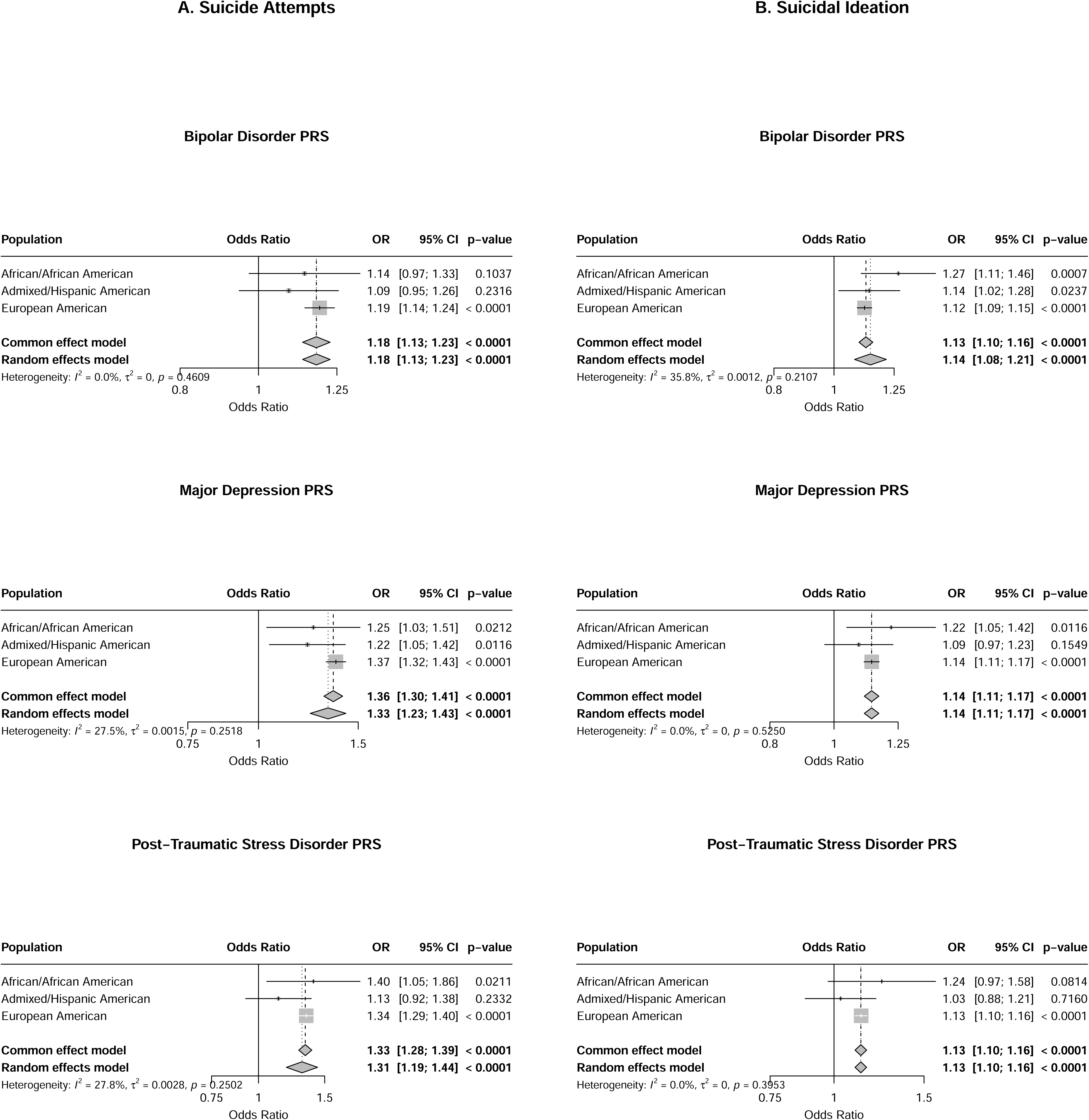
Meta-analyzed Associations Between Polygenic Risk Scores (PRSs) and Suicide-Related Outcomes. Forest plots showing the independent effects of PRSs for depression, bipolar disorder, and PTSD on (A) suicide attempts (SA) and (B) suicidal ideation (SI). Analyses were stratified by ancestry and then meta-analyzed, adjusting for age, sex, socioeconomic covariates, and corresponding psychiatric diagnoses. Each square represents the ancestry-specific odds ratio (OR) with its 95% CI; diamonds denote the meta-analyzed estimate. Both common effect and random effect model-based meta-analysis results are displayed, along with study heterogeneity measures. AFR: African/African American genetic ancestry group, AMR: Admixed/Hispanic American genetic ancestry group, and EUR: European genetic ancestry group.

To assess the robustness of these findings, we conducted multiple sensitivity analyses. Sequential adjustment models showed consistent patterns, with minimally adjusted models that are typically employed in standard PRS analyses yielding up to 13% larger effect sizes than our fully adjusted primary models across PRSs (**eTables 10-11**). When examining psychiatric comorbidity’s influence, PRS associations remained significant even after accounting for both comorbidity and corresponding psychiatric diagnoses (**eTables 12-13**). Varying case and control group definitions (SI cases regardless of SA or restricting controls to individuals without psychiatric disorders) demonstrated similar findings (**eTables 14-15**). Alternative analysis using ordinal regression (**eTables 16-17**) or different PRS construction approaches yielded comparable results (**eTables 18-19**).

### Interactive Effects of Polygenic Risk and Clinical Diagnosis

Last, we examined whether PRS associations with suicide risk varied by clinical diagnosis using interaction analyses. Across ancestries, we found no significant PRS x diagnosis interactions (**eTables 20-21, eFigure 5-6**). As illustrated in Figure 3, marginal effects of PRSs (i.e., slopes) were not statistically significantly different between diagnosed and control groups without psychiatric disorders, although absolute risk remained significantly higher among those with a clinical diagnosis due to their elevated baseline risk (**eTables 22-23**).

**Figure 3.**
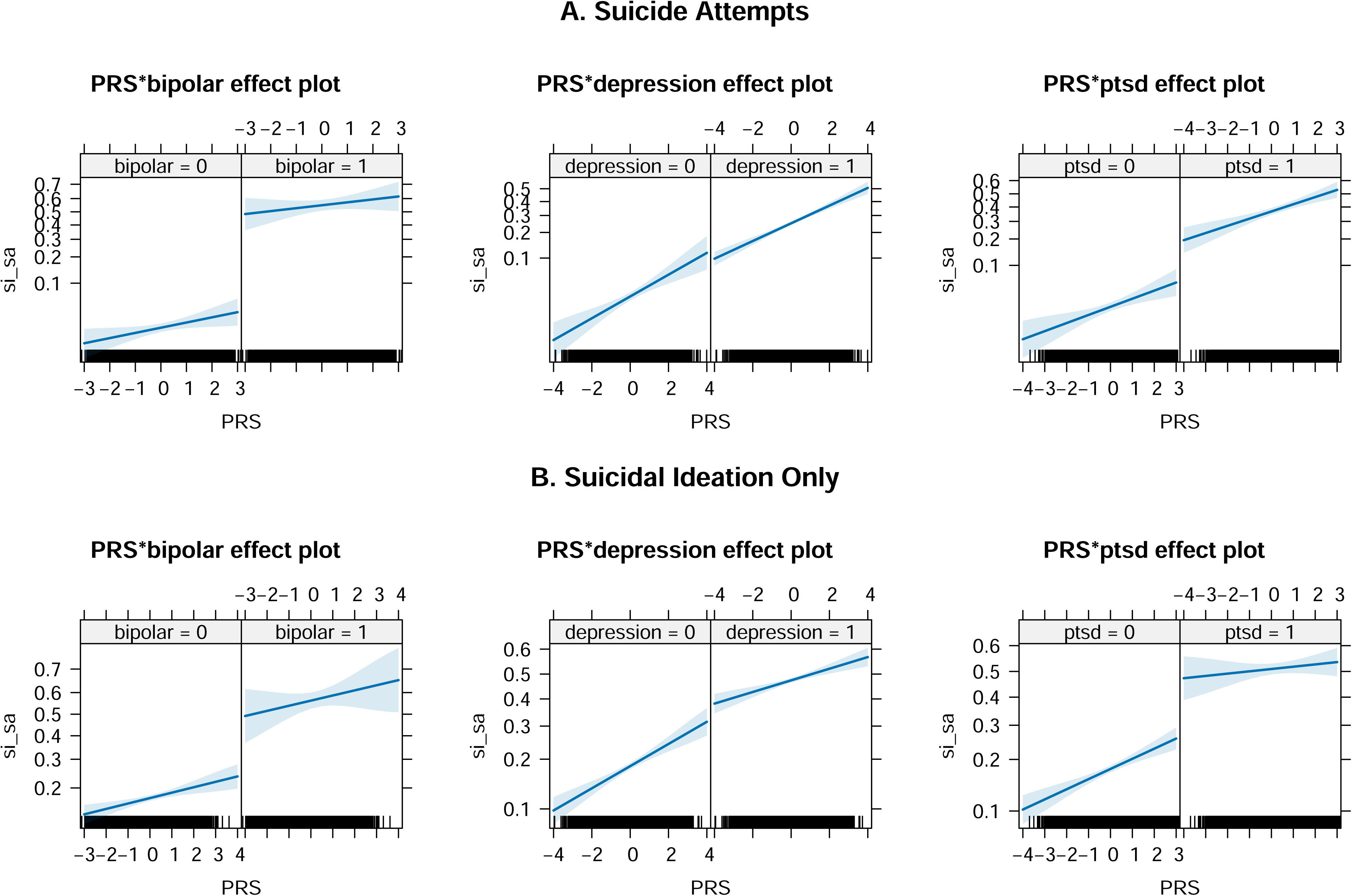
Associations Between Polygenic Risk Scores (PRS) and Suicide-Related Outcomes, Stratified by Psychiatric Diagnosis. (A) Suicide attempt (SA) and (B) suicidal ideation-only (SI) outcome models, showing regression slopes of PRSs stratified by psychiatric diagnoses (PTSD, depression, and bipolar disorder). The Y-axis represents the predicted probability of suicide-related outcomes, while the X-axis shows PRS scores. Separate lines illustrate individuals with and without the corresponding diagnosis, with shaded regions indicating 95% confidence intervals. While PRS effects were not statistically significantly different between diagnostic groups after multiple testing correction at FDR 5% (eTables 16-18), individuals with a psychiatric diagnosis consistently exhibited higher absolute risk than those without the disorder.

## DISCUSSION

Despite extensive research linking psychiatric disorders to suicidal behaviors,^3–7^ current approaches that rely on categorical diagnoses often fail to identify individuals at risk. Recent PRS analyses have shown promise in capturing inherited liability,^10–15,17,22–28^ yet most studies have focused on single disorders, lacked diversity, and rarely evaluated whether genetic risk adds independent value beyond clinical diagnoses. By leveraging the large, ancestrally diverse *All of Us* Research Program,^29–31^ our study extends these important findings by examining multiple psychiatric disorder PRSs across diverse ancestral backgrounds and accounting for a comprehensive range of socioeconomic factors, thereby providing critical new insights into how genetic liability, psychiatric diagnoses, and sociodemographic factors intersect in suicidal thoughts and behaviors.

Consistent with existing literature,^38–42^ we observed strong associations between sociodemographic factors and suicide-related outcomes. Individuals experiencing financial insecurity, living without a partner, identifying with minority sexual orientations, and belonging to certain minoritized racial and ethnic groups showed elevated rates of SI and SA. These findings underscore that suicide risk emerges from the complex interplay of genetic vulnerability and social determinants of health, necessitating integrated prevention approaches that address both biological vulnerabilities and structural and social contexts.

All 13 psychiatric disorders we examined were significantly associated with suicide-related outcomes, after adjusting for age, sex, and socioeconomic factors. Certain disorders such as personality disorders, bipolar disorder, and PTSD showed particularly elevated risk for SA. The cumulative impact of psychiatric comorbidity was striking, with each additional diagnosis more than doubling the odds of SA (OR=2.16 [2.10-2.21]). These findings suggest that cumulative psychiatric burden may be as important as any single diagnosis and reinforce the importance of comprehensive mental health evaluations that go beyond depression screening alone.^43,44^

We also found that PRSs for depression, bipolar disorder, and post-traumatic stress disorder (PTSD) were significantly associated with both SI and SA, independent of sociodemographic factors and corresponding clinical diagnoses. These associations were consistent across individuals with and without lifetime psychiatric diagnoses, highlighting potential transdiagnostic mechanisms of genetic vulnerability. For example, the depression PRS was similarly associated with suicide risk in individuals with (OR=1.31 [1.25-1.37]) and without a depression diagnosis (OR=1.41 [1.25-1.6]). This pattern was observed similarly for PTSD and bipolar disorder PRSs as well, indicating that genetic risk may operate through pathways beyond those reflected in current diagnostic categories.^21,22,45^

Importantly, these findings held across African, Hispanic/Latino, and European ancestry groups. This represents a significant advance, as most prior studies have focused exclusively on individuals of European descent, limiting generalizability. Our results demonstrate that, when derived from well-powered multi-ancestry GWASs,^35–37^ PRSs can show consistent associations with suicide-related outcomes across ancestries. This supports the broader goal of ensuring equity in the application of emerging genomic tools.

From a theoretical standpoint, our findings extend the stress-diathesis model of suicidal behavior by providing empirical evidence for a diathesis that is at least partly genetic, measurable through polygenic scores, and not entirely dependent on the presence of diagnosed psychiatric disorders. This genetic liability may act through intermediate phenotypes, such as affective instability, impaired emotional regulation, impulsivity, or cognitive and behavioral traits that cut across traditional diagnostic boundaries.^46–49^ Future research should investigate how these mechanisms unfold over time and interact with environmental stressors.

Clinically, our findings raise important questions about how to improve suicide risk assessment. Individuals without formal psychiatric diagnoses may still harbor elevated risk due to inherited vulnerability. At the same time, even among those with psychiatric conditions, genetic risk may help explain differences in outcomes. Although current PRSs are not yet ready for clinical use— due to modest effect sizes and ongoing concerns about cross-ancestry portability—they may eventually contribute to multidimensional risk models that integrate genetic, clinical, and environmental information.

Several limitations should be noted. First, the *All of Us* cohort, while diverse, is not nationally representative, ^9,50^ and the opt-in nature of the Emotional Health and Well-Being survey may have introduced selection bias,^11,51^ potentially contributing to the higher observed rates of suicide-related experiences. Second, the cross-sectional nature of our data limit causal inference and precludes temporal analyses of psychiatric diagnoses and suicide-related outcomes. Third, the overlap between SI and core symptoms of major depression may have inflated observed associations. Fourth, while we found no significant PRS-by-diagnosis interactions, our study may have been underpowered to detect such interactions, particularly for outcomes with lower base rates like SA. Finally, our reliance on self-reported lifetime diagnoses may have introduced recall bias.

In summary, our study represents an important step toward precision psychiatry for suicide prevention by demonstrating that genetic vulnerability contributes to suicide risk beyond what is captured by current diagnostic categories and key socioeconomic risk factors across ancestrally diverse populations. These results challenge purely diagnostic models of suicide risk and highlight the need for integrated approaches that combine genetic, clinical, and social dimensions of vulnerability. Future research should aim to refine transdiagnostic risk models, enhance the clinical utility of PRS across diverse populations, and uncover the mechanistic pathways linking genetic predisposition to suicidal behavior.

## Data Availability

All data produced in the present work are contained in the manuscript.

## Author Contributions

PHL and BTS had full access to the *All of Us* data and take responsibility for the integrity and accuracy of the data analysis. PHL and RCK led the conceptualization and design of the study, contributed to data acquisition, analysis, and interpretation, and drafted the manuscript. DYJ, JMG, and BTS were involved in data visualization, statistical analyses, interpretation, and drafting of the manuscript. RCK, RTL, JWS, MKN, JMG, and YHL were involved in critical interpretation of the data and drafting of the manuscript. All authors critically reviewed and participated in the editing of the manuscript for important intellectual content.

## Acknowledgements

We gratefully acknowledge *All of Us* participants for their contributions, without whom this research would not have been possible. We also thank the National Institutes of Health’s *All of Us* Research Program for making available the participant data release (v8) examined in this study. The *All of Us* Research Program is supported by the National Institutes of Health, Office of the Director: Regional Medical Centers: 1 OT2 OD026549; 1 OT2 OD026554; 1 OT2 OD026557; 1 OT2 OD026556; 1 OT2 OD026550; 1 OT2 OD 026552; 1 OT2 OD026553; 1 OT2 OD026548; 1 OT2 OD026551; 1 OT2 OD026555; IAA #: AOD 16037; Federally Qualified Health Centers: HHSN 263201600085U; Data and Research Center: 5 U2C OD023196; Biobank: 1 U24 OD023121; The Participant Center: U24 OD023176; Participant Technology Systems Center: 1 U24 OD023163; Communications and Engagement: 3 OT2 OD023205; 3 OT2 OD023206; and Community Partners: 1 OT2 OD025277; 3 OT2 OD025315; 1 OT2 OD025337; 1 OT2 OD025276. This study also used GWAS summary statistics obtained from the Psychiatric Genomics Consortium (PGC). We would like to thank the research participants and the investigators of these studies for making the data publicly available, which was essential to conducting this study possible.

## Data Sharing Statement

The *All of Us* Research Program data used in this study are available to authorized researchers through the *All of Us* Research website (https://allofus.nih.gov). Detailed information of all outcome and predictor variables obtained from the *All of Us* Research Program data are provided in **eTable 1**. For GWAS summary statistics from the PGC are available to the researchers through the website (https://pgc.unc.edu/for-researchers/download-results/). We provided the detailed download link and study information in **eTable 2**.

## REFERENCES

1. World Health Organization. Suicide. 2025. https://www.who.int/news-room/fact-sheets/detail/suicide

2. CDC. Facts About Suicide. https://www.cdc.gov/suicide/facts/index.html

3. Nock MK, Borges G, Bromet EJ, Cha CB, Kessler RC, Lee S. Suicide and suicidal behavior. Epidemiol Rev. 2008;30(1):133–54. doi:10.1093/epirev/mxn002

4. Franklin J, Ribeiro JD, Fox KR, Bentley KH, Kleiman EM, Huang X, Musacchio KM, Jaroszewski AC, Chang BP, Nock MK. Risk factors for suicidal thoughts and behaviors: A meta-analysis of 50 years of research. Psychol Bul. 2017;143(2):187–232.

5. Cavanagh JT, Carson AJ, Sharpe M, Lawrie SM. Psychological autopsy studies of suicide: a systematic review. Psychol Med. Apr 2003;33(3):395–405. doi:10.1017/s0033291702006943

6. Too LS, Spittal MJ, Bugeja L, Reifels L, Butterworth P, Pirkis J. The association between mental disorders and suicide: A systematic review and meta-analysis of record linkage studies. J Affect Disord. Dec 1 2019;259:302–313. doi:10.1016/j.jad.2019.08.054

7. Mann JJ, Waternaux C, Haas GL, Malone KM. Toward a clinical model of suicidal behavior in psychiatric patients. Am J Psychiatry. Feb 1999;156(2):181–9. doi:10.1176/ajp.156.2.181

8. O’Reilly LM, Pettersson E, Quinn PD, et al. The association between general childhood psychopathology and adolescent suicide attempt and self-harm: A prospective, population-based twin study. Journal of Abnormal Psychology. 2020;129(4):364–375. doi:10.1037/abn0000512

9. Campos A, Verweij KJH, Statham DJ, Madden PAF, Maciejewski DF, Davis KAS, John A, Hotopf M, Heath AC, Martin NG, Rentería ME. Genetic aetiology of self-harm ideation and behaviour. Sci Rep. 2020;10(1):9713.

10. Kimbrel N, Ashley-Koch AE, Qin XJ, Lindquist JH, Garrett ME, Dennis MF, Hair LP, Huffman JE, Jacobson DA, Madduri RK, Trafton JA, Coon H, Docherty AR, Mullins N, Ruderfer DM, Harvey PD, McMahon BH, Oslin DW, Beckham JC, Hauser ER, Hauser MA; Million Veteran Program Suicide Exemplar Workgroup, the International Suicide Genetics Consortium, the Veterans Affairs Mid-Atlantic Mental Illness Research, Education, and Clinical Center Workgroup, and the Veterans Affairs Million Veteran Program. Identification of Novel, Replicable Genetic Risk Loci for Suicidal Thoughts and Behaviors Among US Military Veterans. JAMA Psychiatry. 2023;80(2):135–145.

11. Mullins N, Kang J, Campos AI, Coleman JRI, Edwards AC, et al. Dissecting the shared genetic architecture of suicide attempt, psychiatric disorders and known risk factors. Biol Psychiatry. 2022;91(3):313–327. doi:10.1101/2020.12.01.20241281

12. Kimbrel N, Ashley-Koch AE, Qin XJ, Lindquist JH, Garrett ME, Dennis MF, Hair LP, Huffman JE, Jacobson DA, Madduri RK, Trafton JA, Coon H, Docherty AR, Kang J, Mullins N, Ruderfer DM; VA Million Veteran Program (MVP); MVP Suicide Exemplar Workgroup; International Suicide Genetics Consortium, Harvey PD, McMahon BH, Oslin DW, Hauser ER, Hauser MA, Beckham JC. A genome-wide association study of suicide attempts in the million veterans program identifies evidence of pan-ancestry and ancestry-specific risk loci. Mol Psychiatry. 2022;27(4):2264–2272.

13. Docherty A, Shabalin AA, DiBlasi E, Monson E, Mullins N, Adkins DE, Bacanu SA, Bakian AV, Crowell S, Chen D, Darlington TM, Callor WB, Christensen ED, Gray D, Keeshin B, Klein M, Anderson JS, Jerominski L, Hayward C, Porteous DJ, McIntosh A, Li Q, Coon H. Genome-Wide Association Study of Suicide Death and Polygenic Prediction of Clinical Antecedents. Am J Psychiatry. 2020;177(10):917–927.

14. Mullins N, Bigdeli TB, Borglum AD, et al. GWAS of Suicide Attempt in Psychiatric Disorders and Association With Major Depression Polygenic Risk Scores. Am J Psychiatry. Aug 1 2019;176(8):651–660. doi:10.1176/appi.ajp.2019.18080957

15. Docherty AR, Mullins N, Ashley-Koch AE, et al. GWAS Meta-Analysis of Suicide Attempt: Identification of 12 Genome-Wide Significant Loci and Implication of Genetic Risks for Specific Health Factors. Am J Psychiatry. Oct 1 2023;180(10):723–738. doi:10.1176/appi.ajp.21121266

16. Lewis C, Vassos E. Polygenic risk scores: from research tools to clinical instruments. Genome Med. 2020;12(1):44.

17. Otsuka I, Galfalvy H, Guo J, et al. Relationship of Major Depressive Disorder and Schizophrenia Polygenic Risk Scores to Suicide: A Comparison Between European and Asian Ancestry Populations. Arch Suicide Res. Jan-Mar 2025;29(1):309–316. doi:10.1080/13811118.2024.2332258

18. Lee P, Doyle AE, Silberstein M, Jung J-Y, Liu R, Perlis RH, Roffman J, Smoller JW, Fava M, Kessler RC. Associations between genetic risk for adult suicide attempt and suicidal behaviors in young children: the US population-based study. JAMA Psychiatry. 2022;79(10):971–980.

19. Lee PH, Doyle AE, Li X, et al. Genetic Association of Attention-Deficit/Hyperactivity Disorder and Major Depression With Suicidal Ideation and Attempts in Children: The Adolescent Brain Cognitive Development Study. Biol Psychiatry. Aug 1 2022;92(3):236–245. doi:10.1016/j.biopsych.2021.11.026

20. Lim KX, Rijsdijk F, Hagenaars SP, et al. Studying individual risk factors for self-harm in the UK Biobank: A polygenic scoring and Mendelian randomisation study. PLoS Med. Jun 2020;17(6):e1003137. doi:10.1371/journal.pmed.1003137

21. Stein MB, Jain S, Campbell-Sills L, et al. Polygenic risk for major depression is associated with lifetime suicide attempt in US soldiers independent of personal and parental history of major depression. Am J Med Genet B Neuropsychiatr Genet. Dec 2021;186(8):469–475. doi:10.1002/ajmg.b.32868

22. Stein MB, Jain S, Papini S, et al. Polygenic risk for suicide attempt is associated with lifetime suicide attempt in US soldiers independent of parental risk. J Affect Disord. Apr 15 2024;351:671–682. doi:10.1016/j.jad.2024.01.254

23. Levey DF, Polimanti R, Cheng Z, et al. Genetic associations with suicide attempt severity and genetic overlap with major depression. Transl Psychiatry. Jan 17 2019;9(1):22. doi:10.1038/s41398-018-0340-2

24. Na PJ, De Angelis F, Nichter B, et al. Psychosocial moderators of polygenic risk for suicidal ideation: Results from a 7-year population-based, prospective cohort study of U.S. veterans. Mol Psychiatry. Feb 2022;27(2):1068–1074. doi:10.1038/s41380-021-01352-2

25. Nichter B, Koller D, De Angelis F, et al. Genetic liability to suicidal thoughts and behaviors and risk of suicide attempt in US military veterans: moderating effects of cumulative trauma burden. Psychol Med. Oct 2023;53(13):6325–6333. doi:10.1017/S0033291722003646

26. Campbell-Sills L, Sun X, Papini S, et al. Genetic, environmental, and behavioral correlates of lifetime suicide attempt: Analysis of additive and interactive effects in two cohorts of US Army soldiers. Neuropsychopharmacology. Oct 2023;48(11):1623–1629. doi:10.1038/s41386-023-01596-2

27. Zhang B, You J, Rolls ET, et al. Identifying behaviour-related and physiological risk factors for suicide attempts in the UK Biobank. Nat Hum Behav. Sep 2024;8(9):1784–1797. doi:10.1038/s41562-024-01903-x

28. Ashley-Koch AE, Kimbrel NA, Qin XJ, et al. Genome-wide association study identifies four pan-ancestry loci for suicidal ideation in the Million Veteran Program. PLoS Genet. Mar 2023;19(3):e1010623. doi:10.1371/journal.pgen.1010623

29. All of Us Research Program I, Denny JC, Rutter JL, et al. The "All of Us" Research Program. N Engl J Med. Aug 15 2019;381(7):668–676. doi:10.1056/NEJMsr1809937

30. Bianchi DW, Brennan PF, Chiang MF, et al. The All of Us Research Program is an opportunity to enhance the diversity of US biomedical research. Nat Med. Feb 2024;30(2):330–333. doi:10.1038/s41591-023-02744-3

31. Ramirez A, Sulieman L, Schlueter DJ, Halvorson A, Qian J, Ratsimbazafy F, Loperena R, Mayo K, Basford M, Deflaux N, Muthuraman KN, Natarajan K, Kho A, Xu H, Wilkins C, Anton-Culver H, Boerwinkle E, Cicek M, Clark CR, Cohn E, Ohno-Machado L, Schully SD, Ahmedani BK, Argos M, Cronin RM, O’Donnell C, Fouad M, Goldstein DB, Greenland P, Hebbring SJ, Karlson EW, Khatri P, Korf B, Smoller JW, Sodeke S, Wilbanks J, Hentges J, Mockrin S, Lunt C, Devaney SA, Gebo K, Denny JC, Carroll RJ, Glazer D, Harris PA, Hripcsak G, Philippakis A, Roden DM; All of Us Research Program. The All of Us Research Program: Data quality, utility, and diversity. Patterns (N Y). 2022;3(8):100570.

32. All of Us Research Program Genomics I. Genomic data in the All of Us Research Program. Nature. Mar 2024;627(8003):340–346. doi:10.1038/s41586-023-06957-x

33. Ge T, Chen CY, Ni Y, Feng YA, Smoller JW. Polygenic prediction via Bayesian regression and continuous shrinkage priors. Nat Commun. Apr 16 2019;10(1):1776. doi:10.1038/s41467-019-09718-5

34. Chang CC, Chow CC, Tellier LC, Vattikuti S, Purcell SM, Lee JJ. Second-generation PLINK: rising to the challenge of larger and richer datasets. Gigascience. 2015;4:7. doi:10.1186/s13742-015-0047-8

35. Major Depressive Disorder Working Group of the Psychiatric Genomics Consortium. Electronic address ameau, Major Depressive Disorder Working Group of the Psychiatric Genomics C. Trans-ancestry genome-wide study of depression identifies 697 associations implicating cell types and pharmacotherapies. Cell. Feb 6 2025;188(3):640–652 e9. doi:10.1016/j.cell.2024.12.002

36. O’Connell KS, Koromina M, van der Veen T, et al. Genomics yields biological and phenotypic insights into bipolar disorder. Nature. Jan 22 2025;doi:10.1038/s41586-024-08468-9

37. Nievergelt CM, Maihofer AX, Atkinson EG, et al. Genome-wide association analyses identify 95 risk loci and provide insights into the neurobiology of post-traumatic stress disorder. Nat Genet. May 2024;56(5):792–808. doi:10.1038/s41588-024-01707-9

38. Mpofu JJ, Crosby A, Flynn MA, et al. Preventing Suicidal Behavior Among American Indian and Alaska Native Adolescents and Young Adults. Public Health Rep. Jul-Aug 2023;138(4):593–601. doi:10.1177/00333549221108986

39. Na PJ, Shin J, Kwak HR, et al. Social Determinants of Health and Suicide-Related Outcomes: A Review of Meta-Analyses. JAMA Psychiatry. Jan 2 2025;doi:10.1001/jamapsychiatry.2024.4241

40. Hottes TS, Bogaert L, Rhodes AE, Brennan DJ, Gesink D. Lifetime Prevalence of Suicide Attempts Among Sexual Minority Adults by Study Sampling Strategies: A Systematic Review and Meta-Analysis. Am J Public Health. May 2016;106(5):e1–12. doi:10.2105/AJPH.2016.303088

41. Bridge JA, Asti L, Horowitz LM, et al. Suicide Trends Among Elementary School-Aged Children in the United States From 1993 to 2012. JAMA Pediatr. Jul 2015;169(7):673–7. doi:10.1001/jamapediatrics.2015.0465

42. di Giacomo E, Krausz M, Colmegna F, Aspesi F, Clerici M. Estimating the Risk of Attempted Suicide Among Sexual Minority Youths: A Systematic Review and Meta-analysis. JAMA Pediatr. Dec 1 2018;172(12):1145–1152. doi:10.1001/jamapediatrics.2018.2731

43. Liu RT, Kautz MM, Walsh RFL, Pollak OH, Clayton MG, AR. S. Suicide and depression: Epidemiology, theory, assessment, and treatment. APA Handbook of Depression American Psychological Association. 2025.

44. Nock MK, Borges G, Bromet EJ, et al. Cross-national prevalence and risk factors for suicidal ideation, plans and attempts. Br J Psychiatry. Feb 2008;192(2):98–105. doi:10.1192/bjp.bp.107.040113

45. Kendler KS, Ohlsson H, Moscicki EK, Sundquist J, Edwards AC, Sundquist K. Genetic liability to suicide attempt, suicide death, and psychiatric and substance use disorders on the risk for suicide attempt and suicide death: a Swedish national study. Psychol Med. Mar 2023;53(4):1639–1648. doi:10.1017/S0033291721003354

46. Giner L, Blasco-Fontecilla H, De La Vega D, Courtet P. Cognitive, Emotional, Temperament, and Personality Trait Correlates of Suicidal Behavior. Curr Psychiatry Rep. Nov 2016;18(11):102. doi:10.1007/s11920-016-0742-x

47. Liu RT, Trout ZM, Hernandez EM, Cheek SM, Gerlus N. A behavioral and cognitive neuroscience perspective on impulsivity, suicide, and non-suicidal self-injury: Meta-analysis and recommendations for future research. Neurosci Biobehav Rev. Dec 2017;83:440–450. doi:10.1016/j.neubiorev.2017.09.019

48. Ayer L, Ohana E, Ivanova MY, Frering HE, Achenbach TM, Althoff RR. Emotional and Behavioral Problem Profiles of Preteens With Self-Injurious Thoughts and Behaviors: A Multicultural Study. J Am Acad Child Adolesc Psychiatry. Sep 2024;63(9):931–942. doi:10.1016/j.jaac.2023.11.012

49. Jordan JT, McNiel DE. Characteristics of persons who die on their first suicide attempt: results from the National Violent Death Reporting System. Psychol Med. Jun 2020;50(8):1390–1397. doi:10.1017/S0033291719001375

50. Fry A, Littlejohns TJ, Sudlow C, et al. Comparison of Sociodemographic and Health-Related Characteristics of UK Biobank Participants With Those of the General Population. Am J Epidemiol. Nov 1 2017;186(9):1026–1034. doi:10.1093/aje/kwx246

51. Lee YH, Liu Z, Fatori D, et al. Association of Everyday Discrimination With Depressive Symptoms and Suicidal Ideation During the COVID-19 Pandemic in the All of Us Research Program. JAMA Psychiatry. Sep 1 2022;79(9):898–906. doi:10.1001/jamapsychiatry.2022.1973

