## Supplementary Figure for "Uncovering Genetic Risk Beyond Diagnoses in Suicidal Thoughts and Behaviors: Insights from *All of Us*"

**Supplementary Materials I**

Psychiatric Disorders, Genetic Risk, and Suicidal Thoughts and Behaviors: Insights from the All of Us Research Program

Lee et al. (2025)

• **Supplementary Figures**

eFigure 1. Selection process for study participants in the analysis

eFigure 2. Associations between psychiatric disorders and suicide-related outcomes, adjusting for age, sex, and various socioeconomic factors

eFigure 3. Associations of major depression (MD), post-traumatic stress disorder (PTSD), and bipolar disorder (BIP) PRSs with suicide attempt (SA) outcomes, adjusting for age, sex, SES, and corresponding psychiatric diagnoses

eFigure 4. Associations of MDD, PTSD and BIP PRSs with suicidal ideation (SI) outcomes, adjusting for age, sex, SES, and corresponding psychiatric diagnoses

eFigure 5. PRS x Diagnosis interaction results for suicide attempts (SA)

eFigure 6. PRS x Diagnosis interaction results for suicidal ideation (SI) only

**eFigure 1. Selection process for study participants in the analysis**

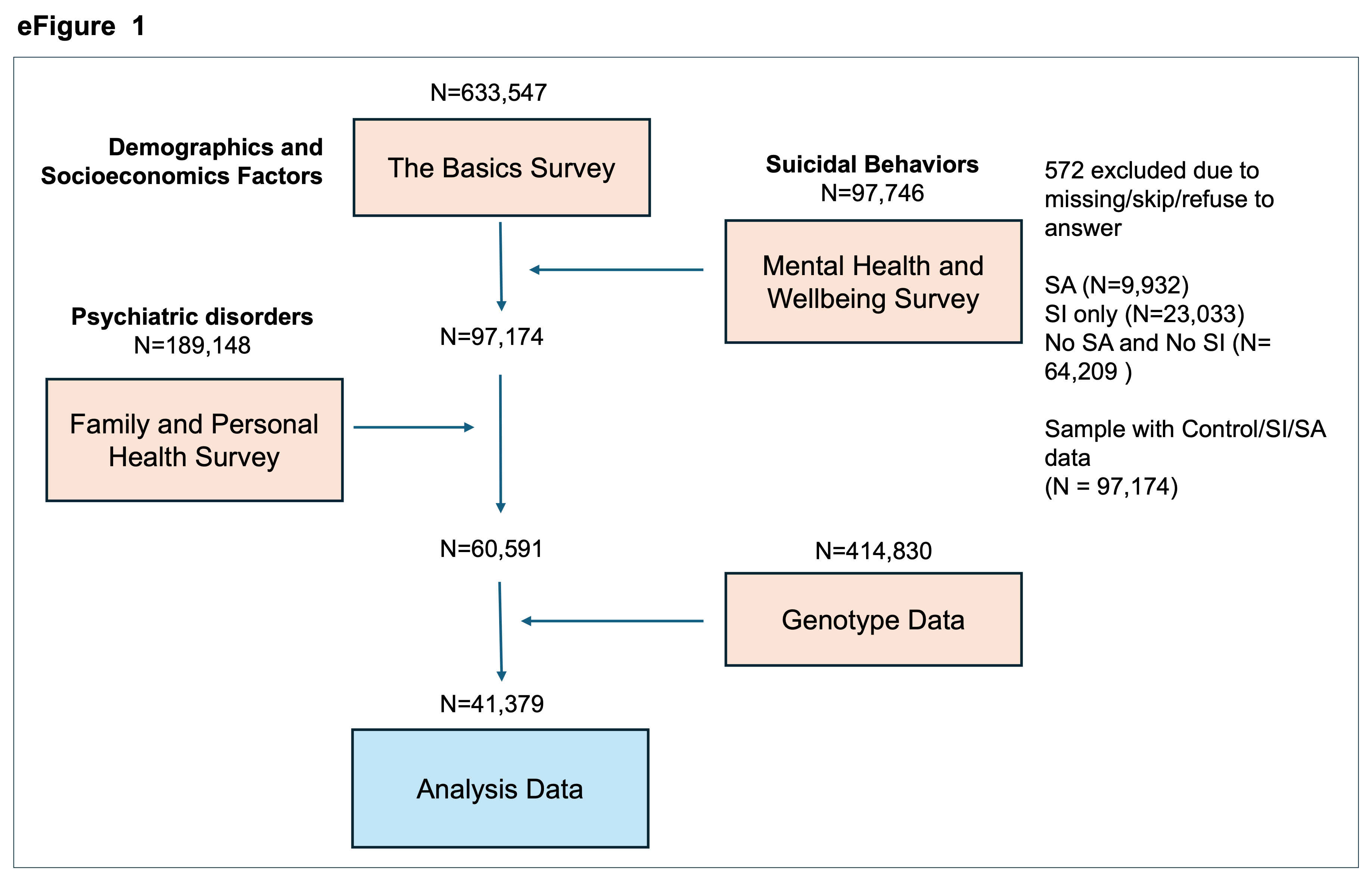

**eFigure 2. Associations between psychiatric disorders and suicide-related outcomes, adjusting for age, sex, and various socioeconomic factors**

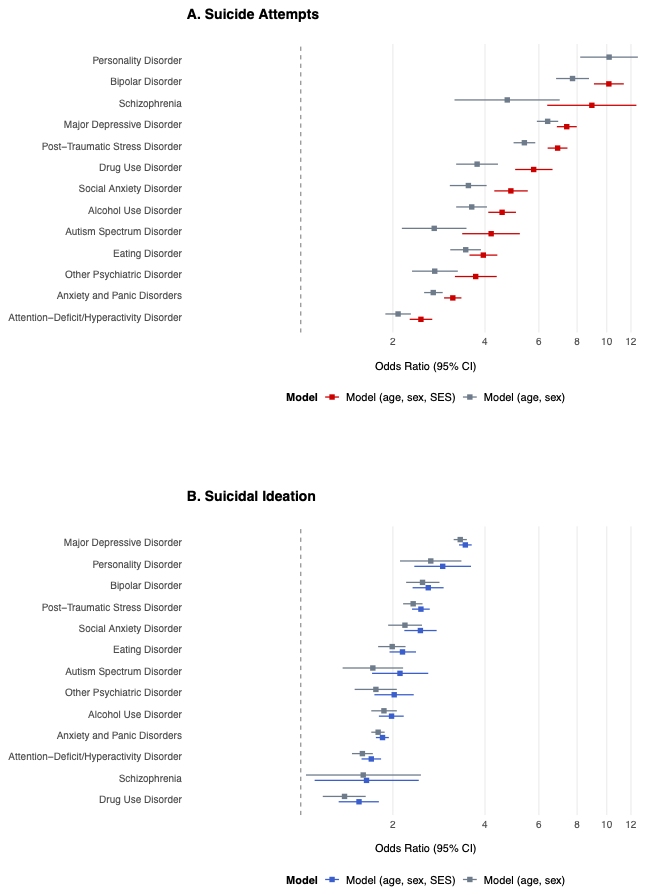

**eFigure 3. Associations of major depression (MD), post-traumatic stress disorder (PTSD), and bipolar disorder (BIP) PRSs with suicide attempt (SA) outcomes, adjusting for age, sex, SES, and corresponding psychiatric diagnoses**

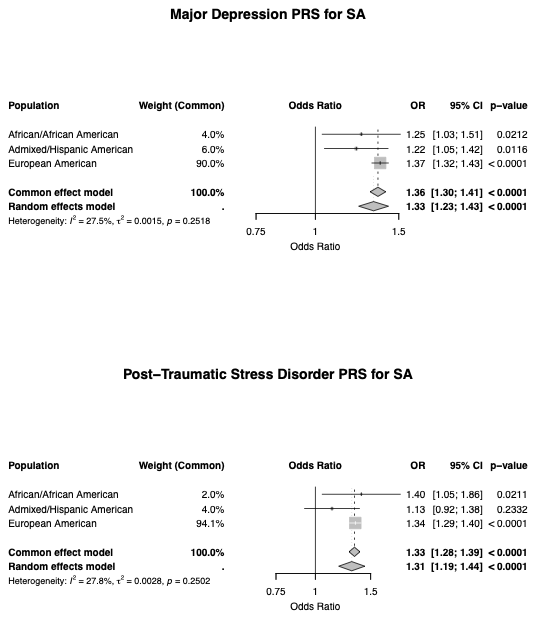

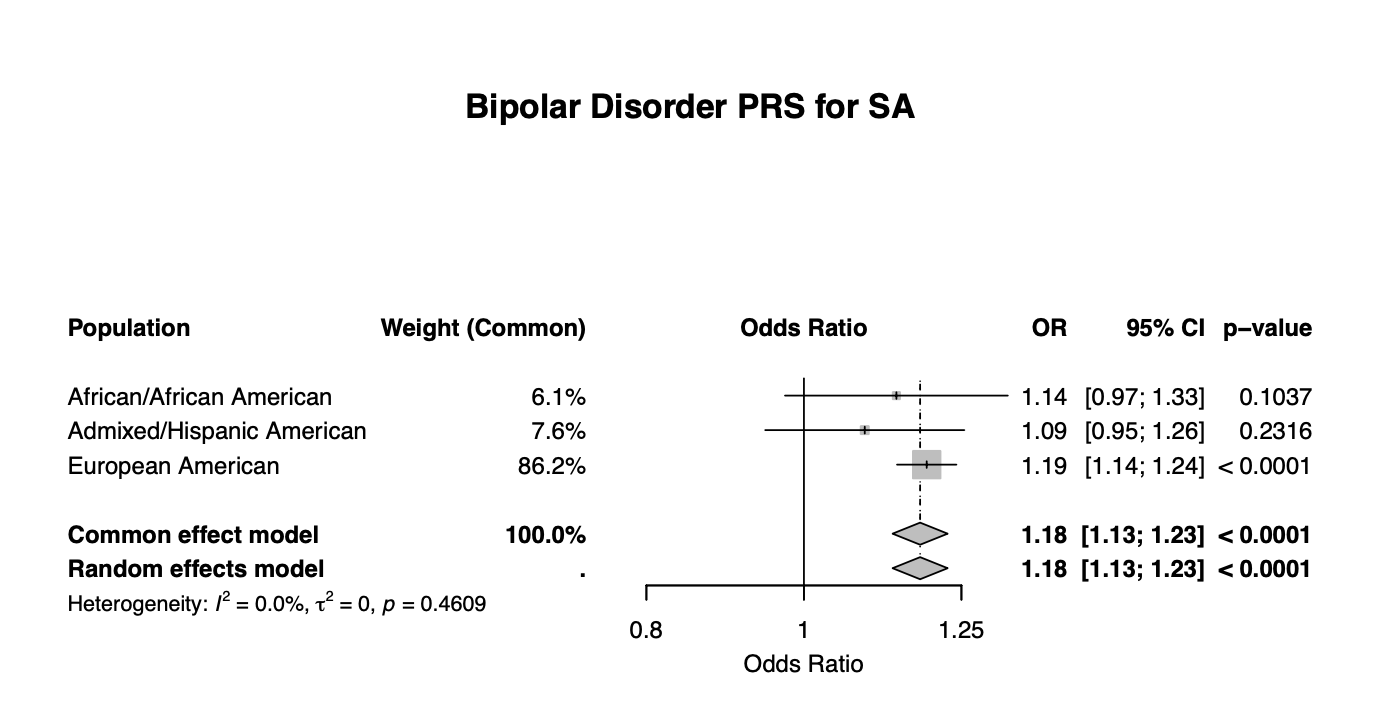

**eFigure 4.** **Associations of MDD, PTSD and BIP PRSs with suicidal ideation (SI) outcomes, adjusting for age, sex, SES, and corresponding psychiatric diagnoses**

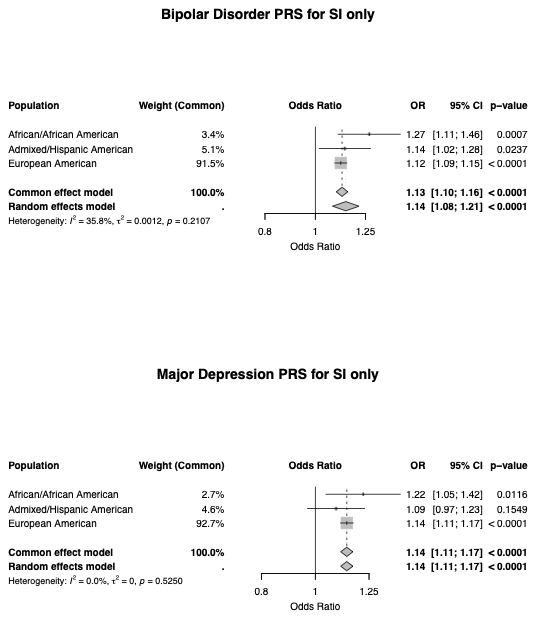

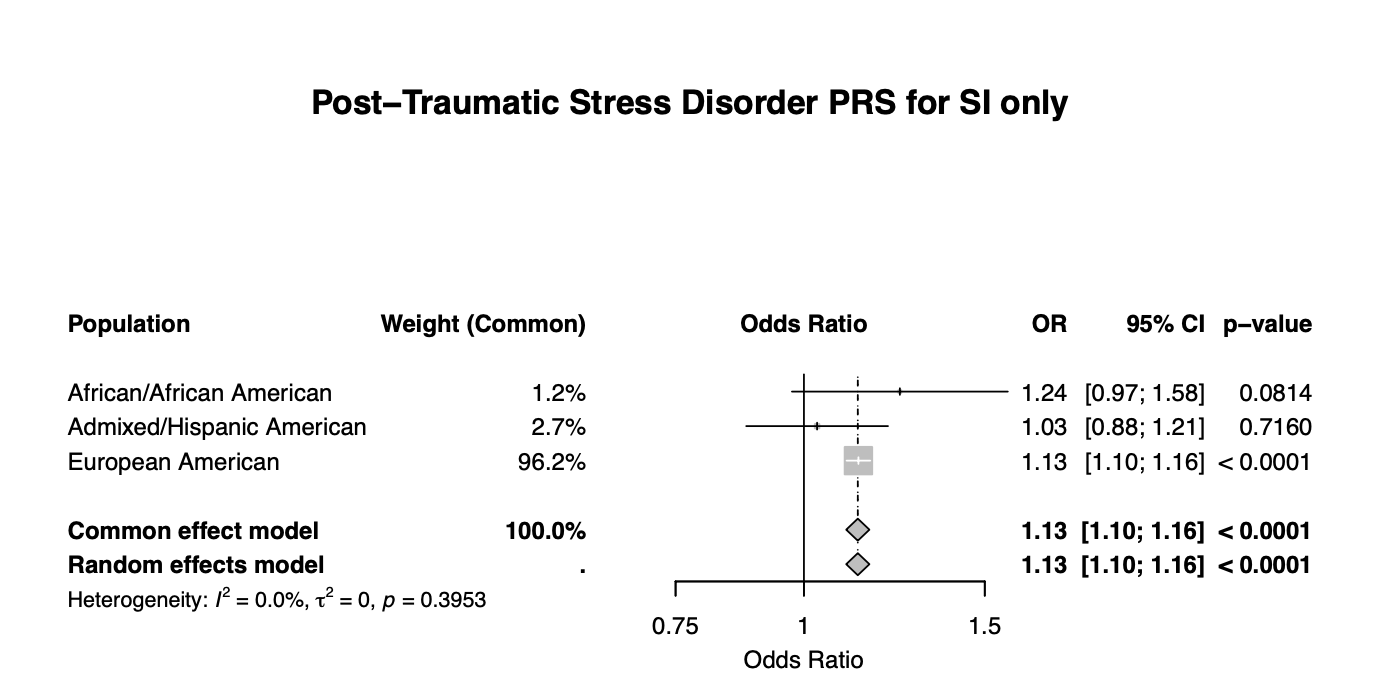

**eFigure 5. PRS x Diagnosis interactions for SA**

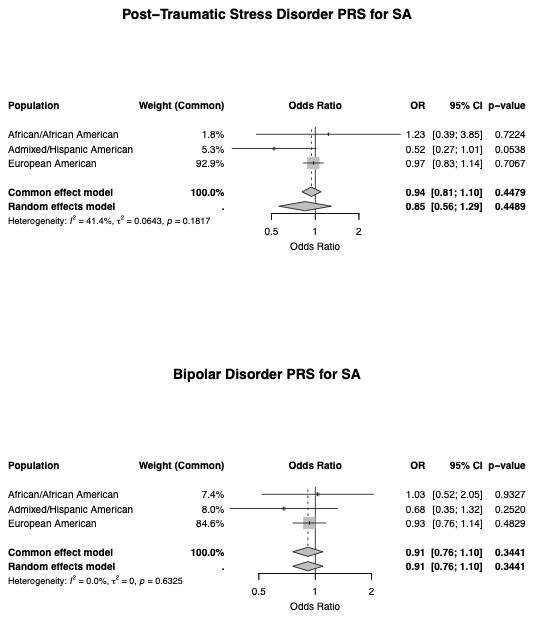

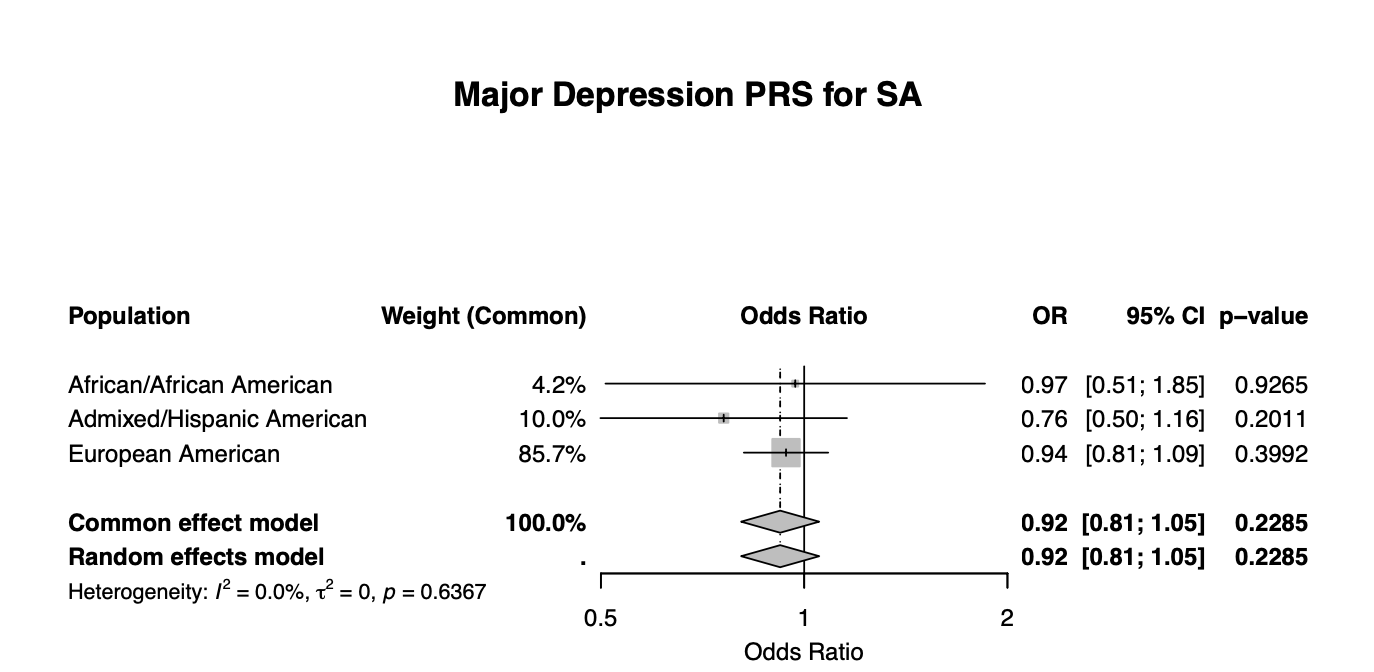

**eFigure 6. PRS x Diagnosis interactions for SI**

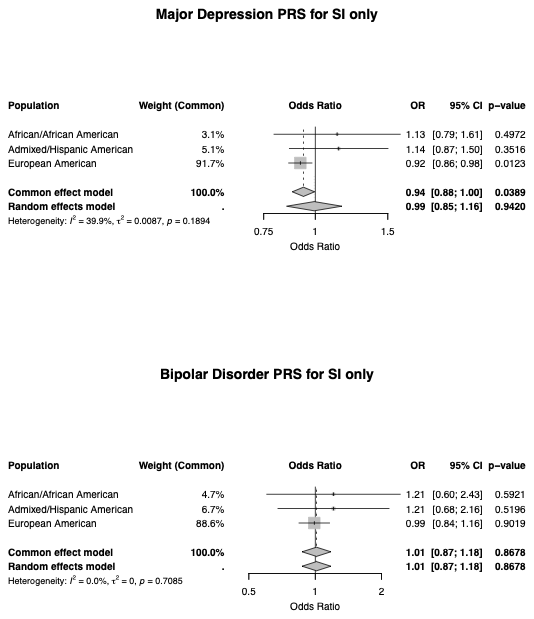

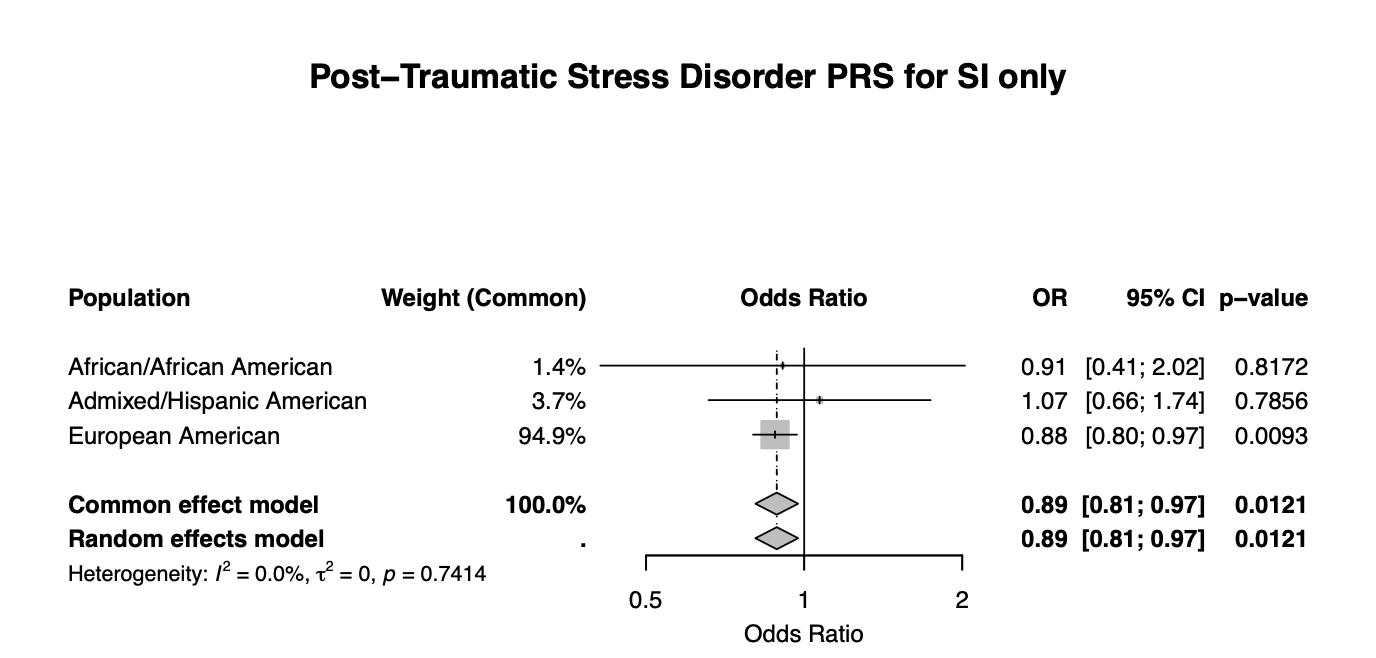
